# Cohen’s kappa statistics as a convenient means to identify accurate SARS-CoV-2 rapid antibody tests

**DOI:** 10.1101/2020.06.13.20130070

**Authors:** Harukazu Iseki

## Abstract

There are many available rapid antibody tests, but the performance of such tests remains unclear. Moreover, it is difficult to compare among the various devices regarding their sensitivity & specificity.

In order to compare the performance of such devices, we used Cohen’ kappa statistics to assess the level of agreement between RT-PCR and rapid antibody tests. In doing this study, we considered the term of validity after symptom-onset to compare two tests. It takes more than a week to produce antibodies in the body, and RT-PCR thus gives negative result in the convalescent period. On ELISA data from the literature kappa statistics was calculated as 1.0 beyond 10 days after symptom-onset. By taking these factors into consideration, we evaluated agreement with samples collected beyond 10 days of symptom-onset during the active period.

We calculated the data from 9 devices, and the kappa statistics for English data were calculated as 0.64 on average. The same finding was 0.75 for Chinese data. These results corresponded with the values from sensitivity & specificity of their reports. Both reports had no details about the collection procedures. Kappa statistics might become even more accurate, if samples could be restricted to ones collected beyond 10 days. Regarding the data from our hospital’, the kappa statistics was 0.97 when restricted to samples collected beyond 10 days, which thus showed excellent agreement.

By using kappa statistics, the performances of rapid antibody tests can be shown as one figure, so that their comparison becomes easy to carry out.

**Highlights:**

▪ Using kappa statistics, agreement between PCR and ELISA was perfect from the data beyond 10 days of symptom-onset.
▪ The results of kappa statistics corresponded with the values from sensitivity & specificity of the English and the Chinese literatures.
▪ Kappa statistics was calculated as 0.97 from the data beyond 10 days of symptom-onset, and sensitivity & specificity were 95.2% and 100% in our hospital.
▪ Kappa statistics is a convenient means to identify accurate rapid antibody tests.

## Introduction

Now the number of new COVID-19 patients has recently begun to decrease. Now several countries want to investigate the SARS-CoV-2 infection rate as an epidemiological study to grasp the actual situation, which will help to evaluate whether or not the acquisition of herd immunity will be possible or not.

There are lots of rapid antibody tests using lateral flow immunoassays method (LFIA). However, their performance remains unclear in large part. It is necessary to estimate the performance of such rapid antibody tests.

It is difficult to judge whether or not a person is infected with COVID-19 under these circumstances, and more than 35% of such patients show poor symptoms ^(1)^, and the sensitivity of reverse transcription-polymerase chain reaction (RT-PCR) assays are reported to be 70% at most.^(2)^ As a result, it is difficult to estimate the agreement between rapid antibody tests and COVID-19 disease in the absence of a reference standard.

We planned to estimate the agreement between rapid antibody tests and RT-PCR using Cohen’s kappa statistics. Kappa (κ) is defined as the difference between the observed and expected agreement expressed as a fraction of the maximum difference and ranges between −1 to 1 ^(3)^.

The accepted data are as follows; “A report from the National COVID Scientific Advisory Panel” (England) ^(4)^, “Application of A Rapid IgM-IgG Combined Antibody Test for SARS-CoV-2” (China) ^(5)^, and Sagamihara Kyodo Hospital ‘s trial (Japan).

## Methods

Agreements between PCR and rapid antibody tests were calculated using the kappa statistics on samples from the English literature, the Chinese literature and our hospital’s patients, who were suspected to have COVID-19 from late April 2020.

We could also accept results using Enzyme-linked immunosorbent assay (ELISA) data in an English report. Kappa statistics was calculated as 1.0 between PCR and ELISA when restricted to samples collected beyond 10 days of symptom-onset. It takes more than a week to produce antibodies in the body. In addition, the PCR reaction becomes negative during the convalescent period.

Judging by the results of this survey, we concluded that samples should be collected beyond 10 days of symptom-onset until the end of the active period as an appropriate period for the agreement between PCR and rapid antibody tests. We regret details regarding the timing of the collected samples from both reports were unknown. For our hospital data, however, we could evaluate the agreement between PCR and rapid antibody test during the appropriate period.

### Statistical Analysis

|  | PCR(+) | PCR(-) |
| --- | --- | --- |
| IgG(+) | a | b |
| IgG(-) | c | d |

## Results

1. In an English report (Table I & Table II) Agreement between PCR and ELISA was evaluated using kappa statistics. Kappa statistics was calculated as 0.86±0.03 from the overall data. On the other hand, kappa statistics was 1.00 from the data collected beyond 10 days of symptom-onset. (Table I). Agreement between PCR and antibody tests from 9 devices was evaluated using kappa statistics. The average value was 0.64±0.03. The details are shown in the Table II.
2. In a Chinese report (Table III) Agreement between PCR and overall 434 samples of antibody tests were evaluated. Kappa statistics was calculated as 0.75.
3. For our hospital’s data (Japan) (Table IV) In the overall data, sensitivity was 0.84, specificity was 0.97, and kappa statistics was calculated as 0.83±0.04. On the other hand, kappa statistics was 0.97±0.02 from the data collected beyond 10 days of symptom-onset.

**[Table I].**
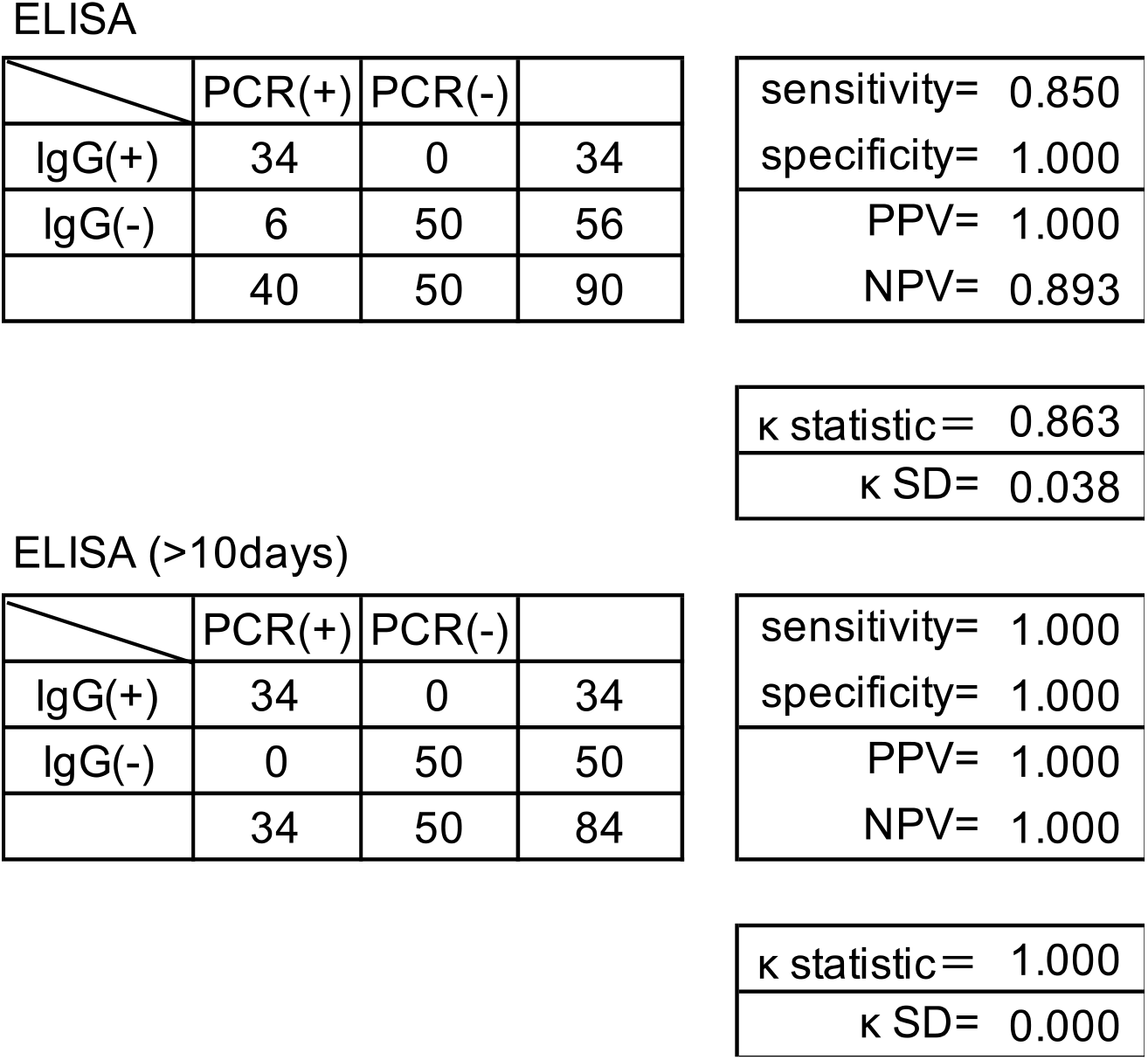
Agreement between PCR and ELISA in an English report using kappa statistics The table above shows the result calculated from the overall data. The table below shows the result calculated from the data collected beyond 10 days of symptom-onset. (PPV: positive predictive value, NPV: negative predictive value)

|  | PCR(+) | PCR(-) |  |
| --- | --- | --- | --- |
| IgG(+) | 34 | 0 | 34 |
| IgG(-) | 6 | 50 | 56 |
|  | 40 | 50 | 90 |

|  |
| --- |
| sensitivity= 0.850 |
| specificity= 1.000 |
| PPV= 1.000 |
| NPV= 0.893 |
| $\kappa$ statistic= 0.863 |
| $\kappa$ SD= 0.038 |

ELISA (>10days)
|  | PCR(+) | PCR(-) |  |
| --- | --- | --- | --- |
| IgG(+) | 34 | 0 | 34 |
| IgG(-) | 0 | 50 | 50 |
|  | 34 | 50 | 84 |

|  |
| --- |
| sensitivity= 1.000 |
| specificity= 1.000 |
| PPV= 1.000 |
| NPV= 1.000 |
| $\kappa$ statistic= 1.000 |
| $\kappa$ SD= 0.000 |

**[Table II].**
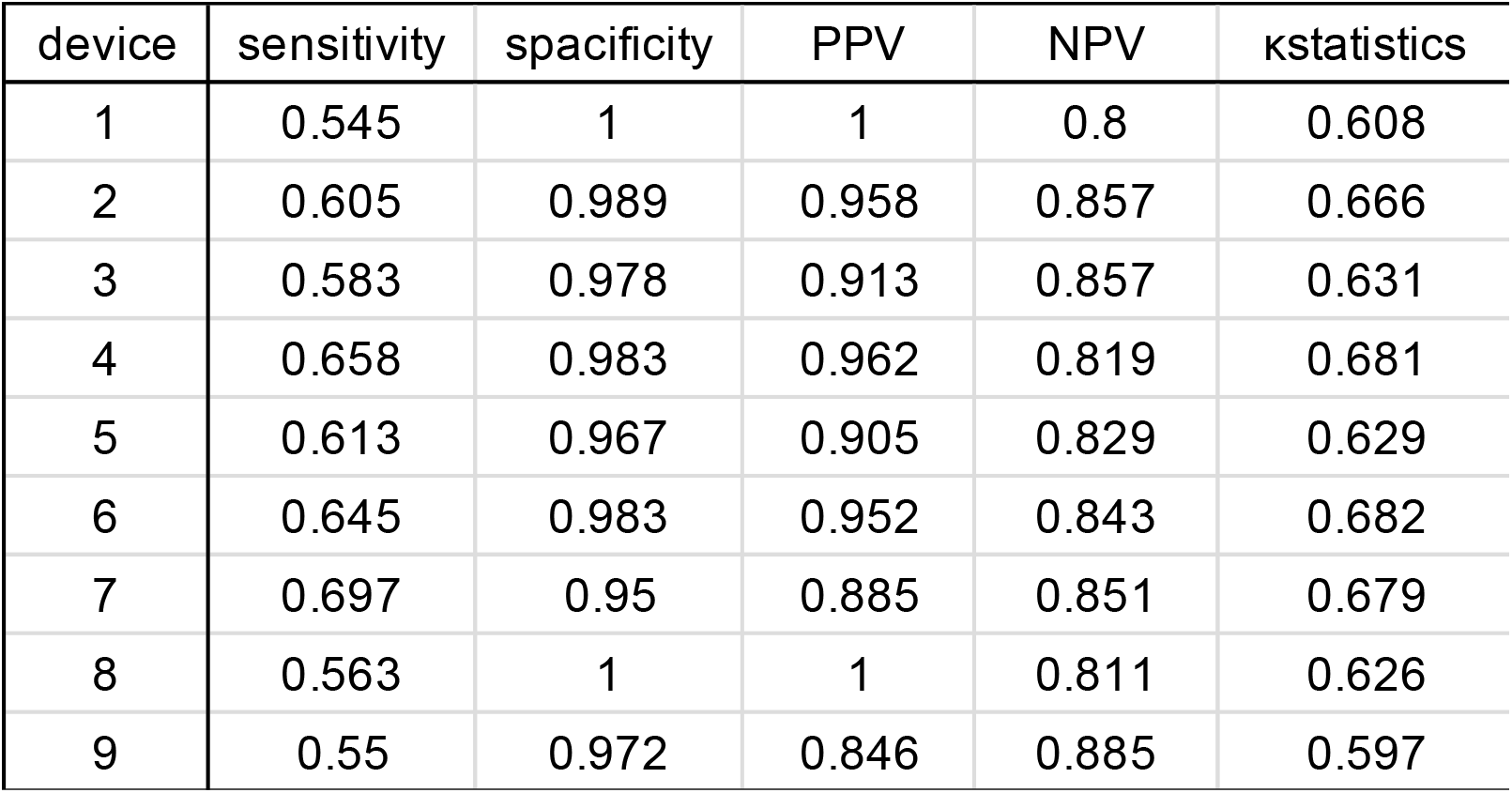
Agreement between PCR and rapid antibody testing from 9 devices in an English report using kappa statistics (PPV: positive predictive value, NPV: negative predictive value)

**[Table III].**
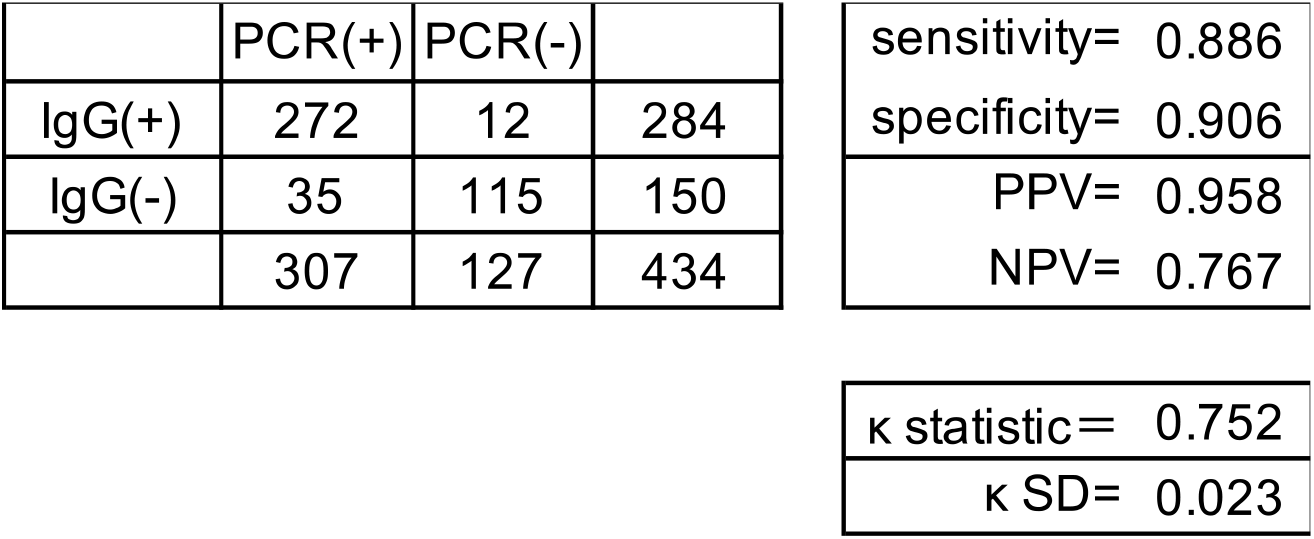
Agreement between PCR and rapid antibody testing in a Chinese report using kappa statistics.

|  | PCR(+) | PCR(-) |  | sensitivity= 0.886 |
| --- | --- | --- | --- | --- |
| IgG(+) | 272 | 12 | 284 | specificity= 0.906 |
| IgG(-) | 35 | 115 | 150 | PPV= 0.958 |
|  | 307 | 127 | 434 | NPV= 0.767 |

|  |
| --- |
| $\kappa$ statistic= 0.752 |
| $\kappa$ SD= 0.023 |

**[Table IV].**
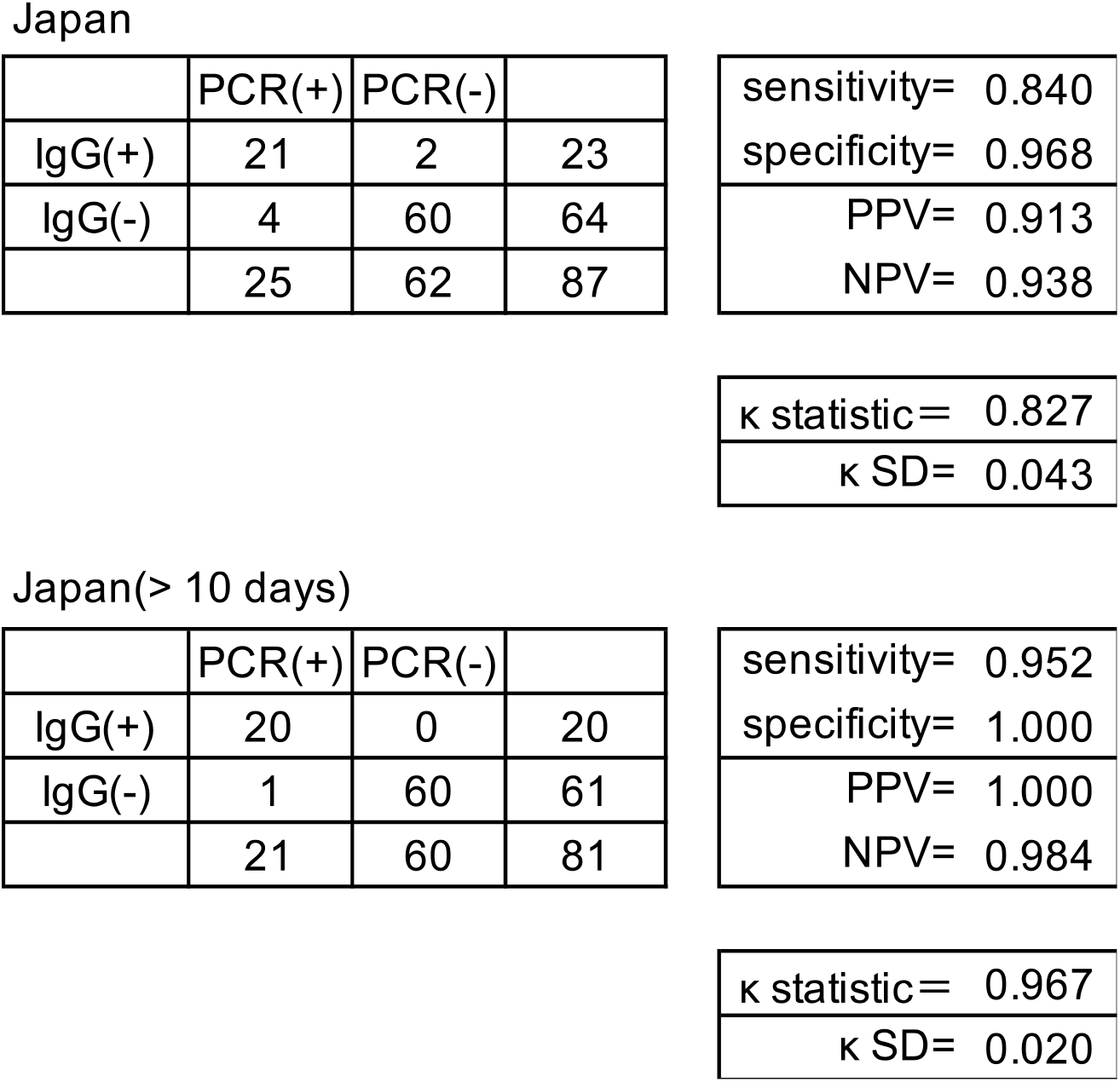
Agreement between PCR and rapid antibody testing in our hospital using kappa statistics The table above shows the result calculated from the overall data. The table below shows the result calculated from the data collected beyond 10 days of symptom-onset.

|  | PCR(+) | PCR(-) |  | sensitivity= 0.840 |
| --- | --- | --- | --- | --- |
| IgG(+) | 21 | 2 | 23 | specificity= 0.968 |
| IgG(-) | 4 | 60 | 64 | PPV= 0.913 |
|  | 25 | 62 | 87 | NPV= 0.938 |

|  |
| --- |
| $\kappa$ statistic= 0.827 |
| $\kappa$ SD= 0.043 |

Japan(> 10 days)
|  | PCR(+) | PCR(-) |  | sensitivity= 0.952 |
| --- | --- | --- | --- | --- |
| IgG(+) | 20 | 0 | 20 | specificity= 1.000 |
| IgG(-) | 1 | 60 | 61 | PPV= 1.000 |
|  | 21 | 60 | 81 | NPV= 0.984 |

|  |
| --- |
| $\kappa$ statistic= 0.967 |
| $\kappa$ SD= 0.020 |

Sensitivity and specificity were 95.2% and 100% when restricted to these data. About our excluded data

1. PCR(+)IgG(-): 3samples within 10 days of symptom-onset
2. PCR(+)IgG(+): one sample collected at the 8^th^day of symptom-onset
3. PCR(-)IgG(+): a patient suffered from an unknown type of pneumonia in March. The patient was in the convalescent period

## Discussion

PCR shows a positive finding early in the active period, but it becomes negative in the convalescent period. On the other hand, antibody test shows a negative finding early in the active period because it takes more than a week to produce antibodies in the body.

In an Oxford study, ELISA data show all samples positive beyond 10 days of symptom-onset. Base on this result, samples beyond 10 days in the active period were accepted for assessing agreement in the absence of a reference standard.

Two reports did not show the details of samples. It would be better if samples could be narrowed down to those collected beyond 10 days.

In our hospital’s data, we calculated kappa statistics using the overall data at first, which was 0.83±0.04.

Then we calculated kappa statistics for samples beyond 10 days, which was 0.97±0.02. Sensitivity was 95.2% and specificity was 100%. These were considered to be excellent results.

Our study shows a simple evaluation of antibody tests using kappa statistics between PCR and antibody tests in the appropriate period for the collection of samples.

Regarding limitations associated with this study, our positive data could not be collected as well as expected because SARS-CoV-2 infection rate has recently decreased, and we could not collect as many samples as we had expected.

From our study, however, we believe the proper antibody test kits can be identified using kappa statistics.

## Data Availability

All data referred to in the manuscript are included in this published article.

## ACKNOWLEDGMENTS

We wish to thanks Mr. Kenichi Kaneko, (President & CEO of LOKI CONSULTING Co., Ltd.), for supporting rapid antibody test devices at no charge.

